# Comparing surveyed adults with long COVID and those with just a positive test helps put COVID into perspective

**DOI:** 10.1101/2024.02.02.24302216

**Authors:** Mary L. Adams, Joseph Grandpre

## Abstract

**Introduction:** Among adults who test positive for COVID-19, some develop long COVID (symptoms lasting ≥3 months), and some do not. We compared 3 groups on selected measures to help determine strategies to reduce COVID impact.

**Methods:** Using Stata and data for 385,617 adults from the 2022 Behavioral Risk Factor Surveillance System, we compared adults reporting long COVID, those with just a positive test, and those who never tested positive, on several health status and risk factor measures plus vaccination rates (data for 178,949 adults in 29 states).

**Results:** Prevalence of just COVID was 26.5% (95% CI 26.2-26.8) and long COVID was 7.4% (7.3-7.6). Compared with adults with just COVID those with long COVID had worse rates for 13 of 17 measures of chronic disease, disability, and poor health status, while those with just COVID had the best results for 15 of the 17 measures among all 3 groups. The 5 risk factors (obesity, diabetes, asthma, cardiovascular disease, and COPD) previously associated with COVID deaths, increased long COVID but not just COVID rates, which were highest among younger and higher income adults. Adults with long COVID had the highest rate among the 3 groups for any COVID risk factors and data from 29 states showed they had the lowest rates for ≥3 vaccine doses of 35.6%, vs. 42.7% and 50.3% for those with just a positive test, and neither, respectively. Vaccination with ≥3 vaccines vs. <3 reduced long COVID rates by 38%, and just COVID rates by 16%.

**Conclusions:** Results show the seriousness of long COVID vs. just a positive test and that increasing vaccine coverage by targeting adults with risk factors shows promise for reducing COVID impact.

## Introduction

Some adults who test positive for COVID-19 develop long COVID while most do not. Long Covid can cause a wide range of ongoing symptoms in people who once had COVID-19 and can be considered a disability under the Americans with Disabilities Act [1]. The Behavioral Risk Factor Surveillance System (BRFSS) is an ongoing state-based telephone survey of randomly selected non-institutionalized adults that collects data on a range of health topics [2]. Results from the 2022 BRFSS on the prevalence of ever having a positive test for COVID have been reported for all participating states and territories [3] and limited to the 50 states and DC [4] with both studies finding a rate of 34.4%. Only the study based on the 50 states [4] reported a rate for long COVID using the definition of symptoms lasting ≥3 months as a percentage of all adults; that rate was 7.4% (95% CI 7.3-7.6). The other study reported long COVID as a percentage of adults with a positive test, which was 21.8% [3]. This study was undertaken to learn more about the similarities and differences between adults with long COVID and those with just a positive test and those never testing positive. The BRFSS provides an opportunity to compare rates with other survey measures of disability, health status, chronic diseases, health care access, and risk factors to help put the potential burden of COVID and long COVID into perspective. Because most data are available for all 50 states, regional and state differences could be considered. The 2022 BRFSS also included optional questions on COVID vaccinations that could provide potentially useful information for 29 states.

Our main study objective was to compare adults with long COVID (symptoms lasting ≥3 months), those having a positive test that did not develop into long COVID (just COVID), and those never testing positive, on a broad range of disability and health status measures. Our second objective was to determine vaccination rates and approximate effectiveness of vaccines on reducing rates of long COVID and just COVID using data from the 29 states that included optional questions on COVID vaccinations.

## Methods

BRFSS data are publicly available from all 50 states and DC along with survey questions and information needed for analysis on the Centers for Disease Control and Prevention (CDC) website [5]. Data were already weighted to adjust for the probability of selection and to reflect the adult population of each state by age group, race/ethnicity, education level, marital status, and home ownership. The median response rate for the 50 states plus DC for land line and cell phone surveys combined was 45.1% [5], ranging from 36.2% to 66.8%. A total of 385,617 respondents were included in most analysis with state N’s ranging from 3,188 in NV to 26,152 in WA and a median N of 7,473.

Measures: Long COVID was defined as ever testing positive for COVID and reporting symptoms lasting 3 months or longer. Those testing positive without long COVID were considered to have just COVID. Respondents with long COVID were asked about the primary symptom they experienced, with a list read that included “no long-term symptoms that limited your activities.” Other measures included age, race/ethnicity, gender, income, education, children in the household, health insurance, employment, disability status (difficulty with hearing, seeing, cognition, walking, dressing, or doing errands alone), frequent (≥14) days of activity limitation in past month (FAL), frequent poor mental health days (FMD), fair or poor general health vs good or better, a depression diagnosis, census region (Northeast, Midwest, South and West), blue vs. red states based on voting in the 2020 Presidential election, current smoking, e-cigarette use, and any HIV risk factor in past year (injected drug use, STD treatment, or exchanging sex for money or drugs, had anal sex without a condom, or had four or more sex partners)[4, 5]. A composite measure of 5 chronic conditions (obesity, asthma, cardiovascular disease (CVD), diabetes, and Chronic Obstructive Pulmonary Disease (COPD) among 6 found to be associated with US hospitalizations for COVID [6,7] was included. Data on hypertension, the 6^th^ chronic condition, was not available for 2022.

Analysis: Stata version 18.0 (StataCorp LLC, College Station TX) was used to account for the complex sample design of the BRFSS in unadjusted analysis and controlled for the listed factors in multiple logistic regression. The survey measures used to describe the survey design were_psu and _ststr, weight=_llcpwt; linearized variance estimation was selected, and for strata with a single sampling unit, center at the grand mean. Missing values (responses of “don’t know” or refusals) for any measure except income were excluded from analysis. Separate univariate analysis was done for just COVID and long COVID (Table 1) to determine the prevalence rates of each for different groups and to help select variables to be included in logistic regression models. Measures with adjusted odds ratios (AORs) having P values >0.05 were removed and only the final logistic regression models are reported. A 3-level measure with the mutually exclusive categories of just COVID, long COVID, and neither was used to compare the groups on a range of health and disability measures (Table 3). Those results compare the rates of the various health measures for adults in each of the 3 groups.

**Table 1.**
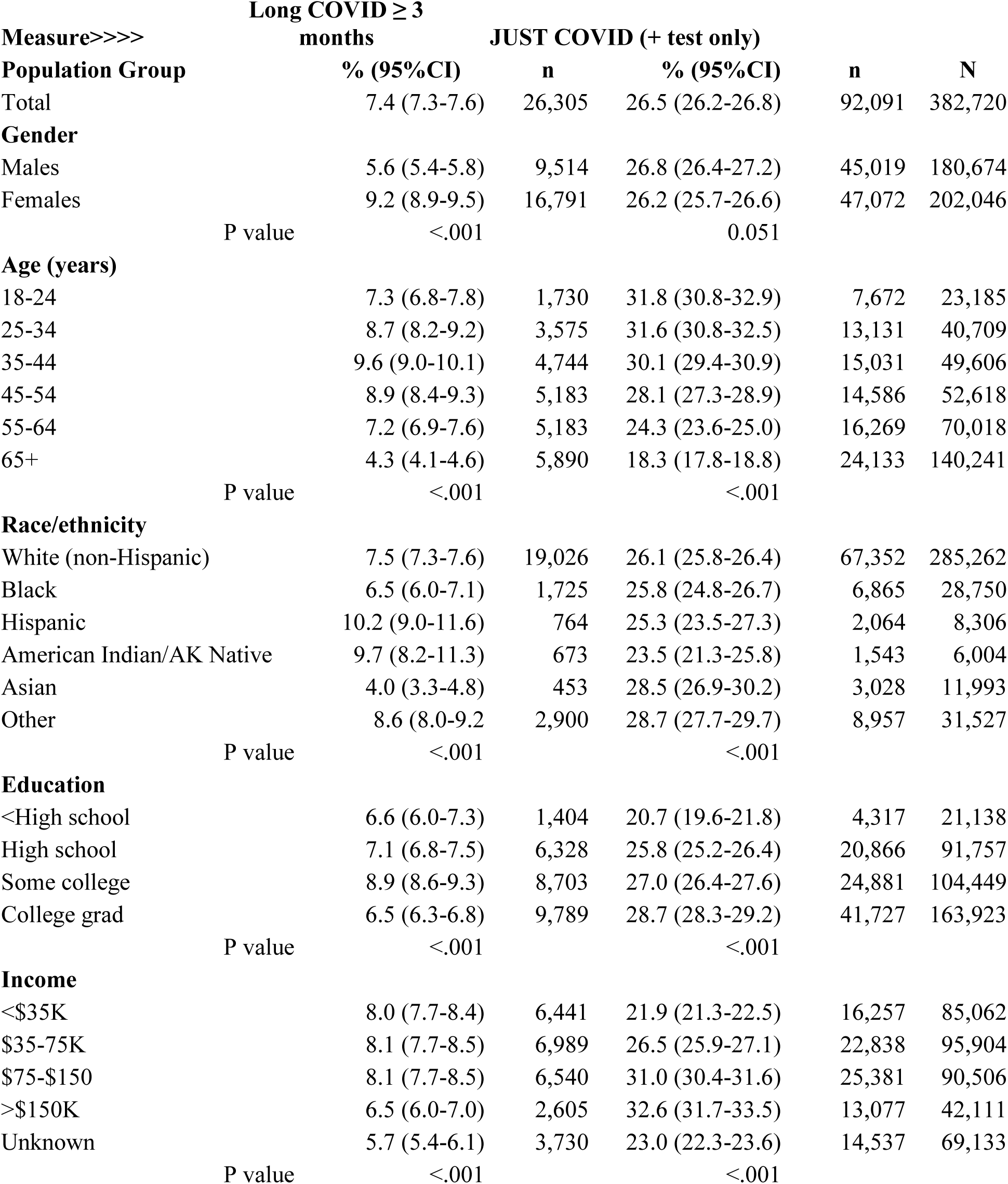
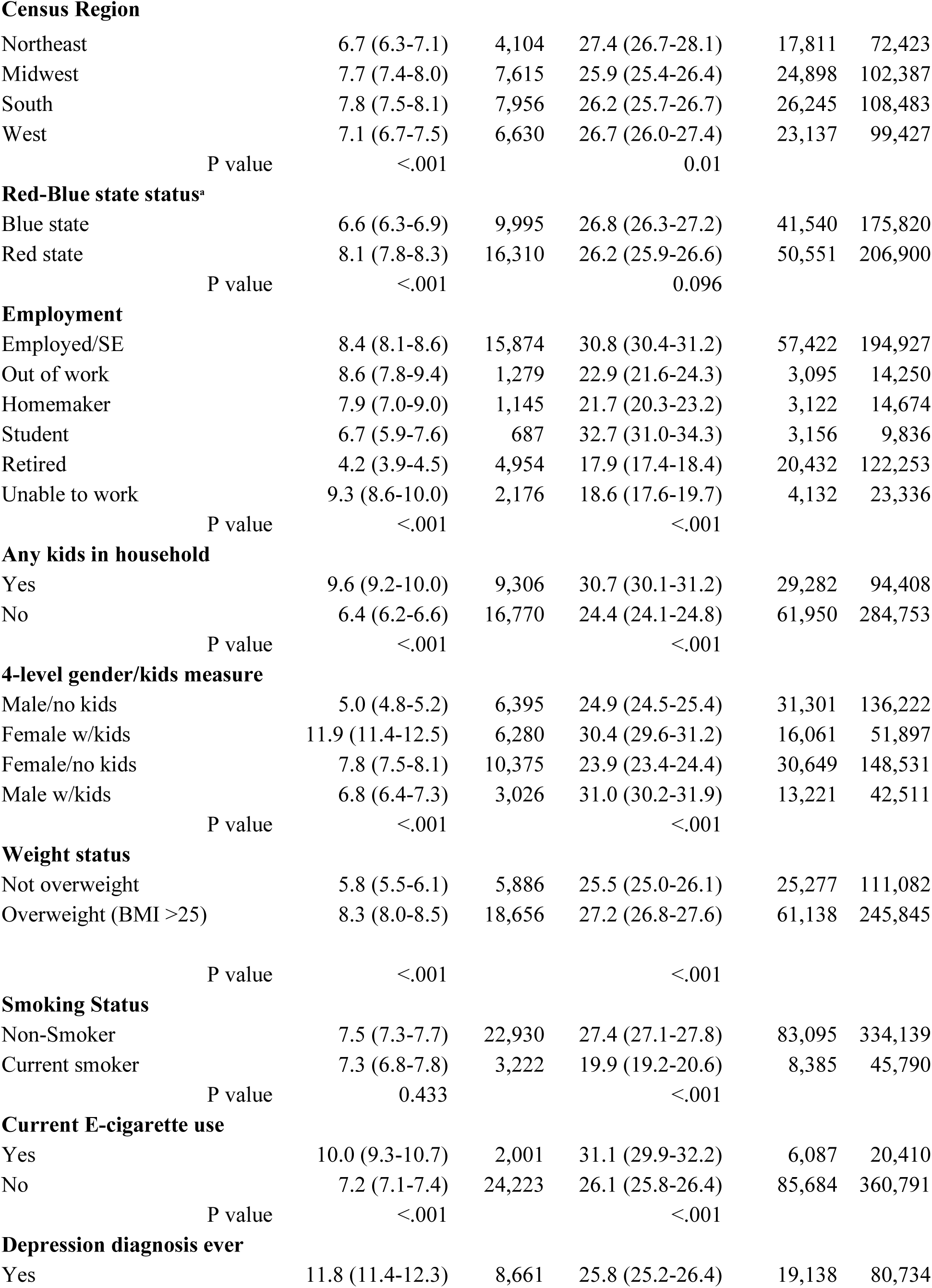
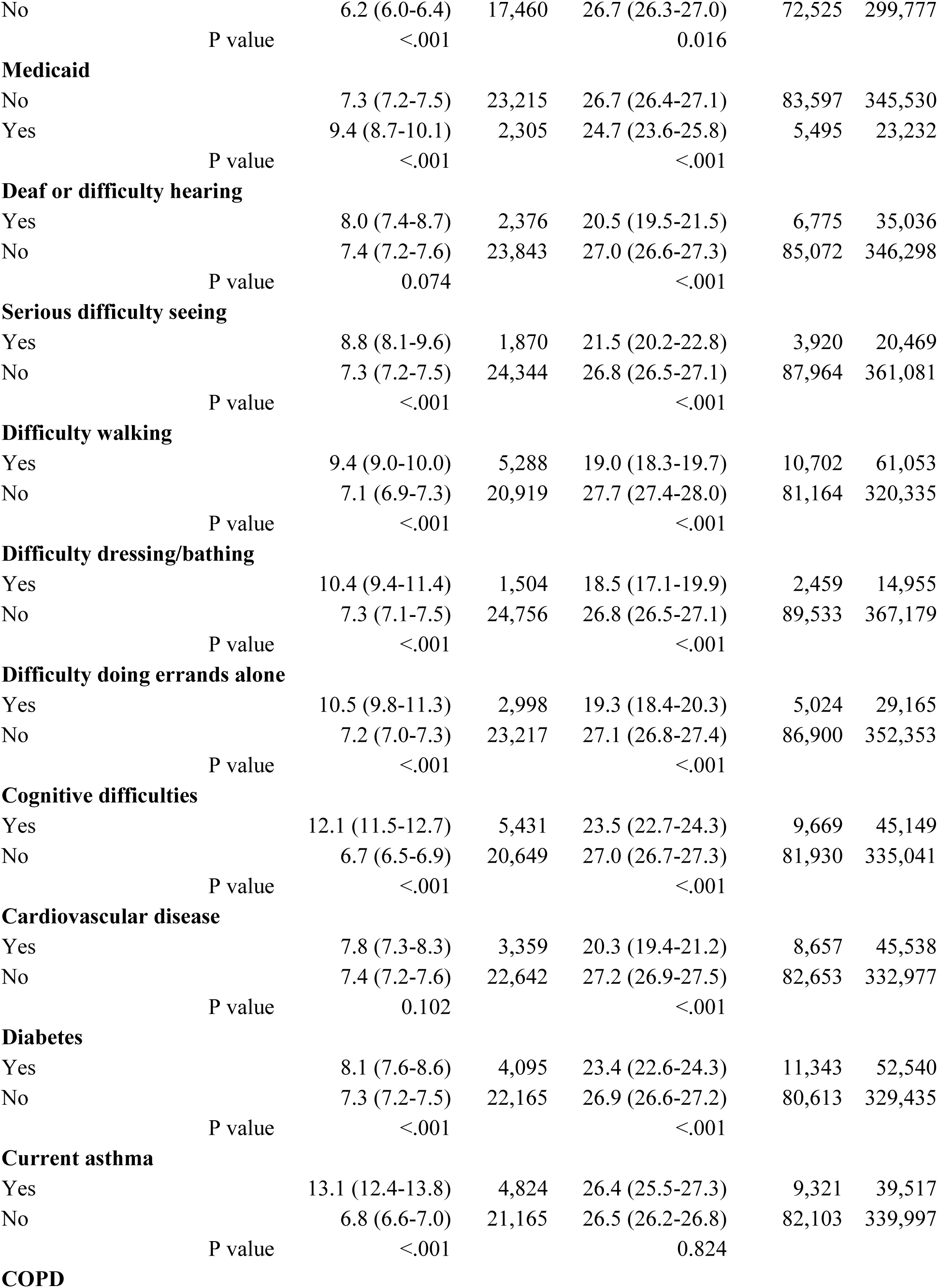
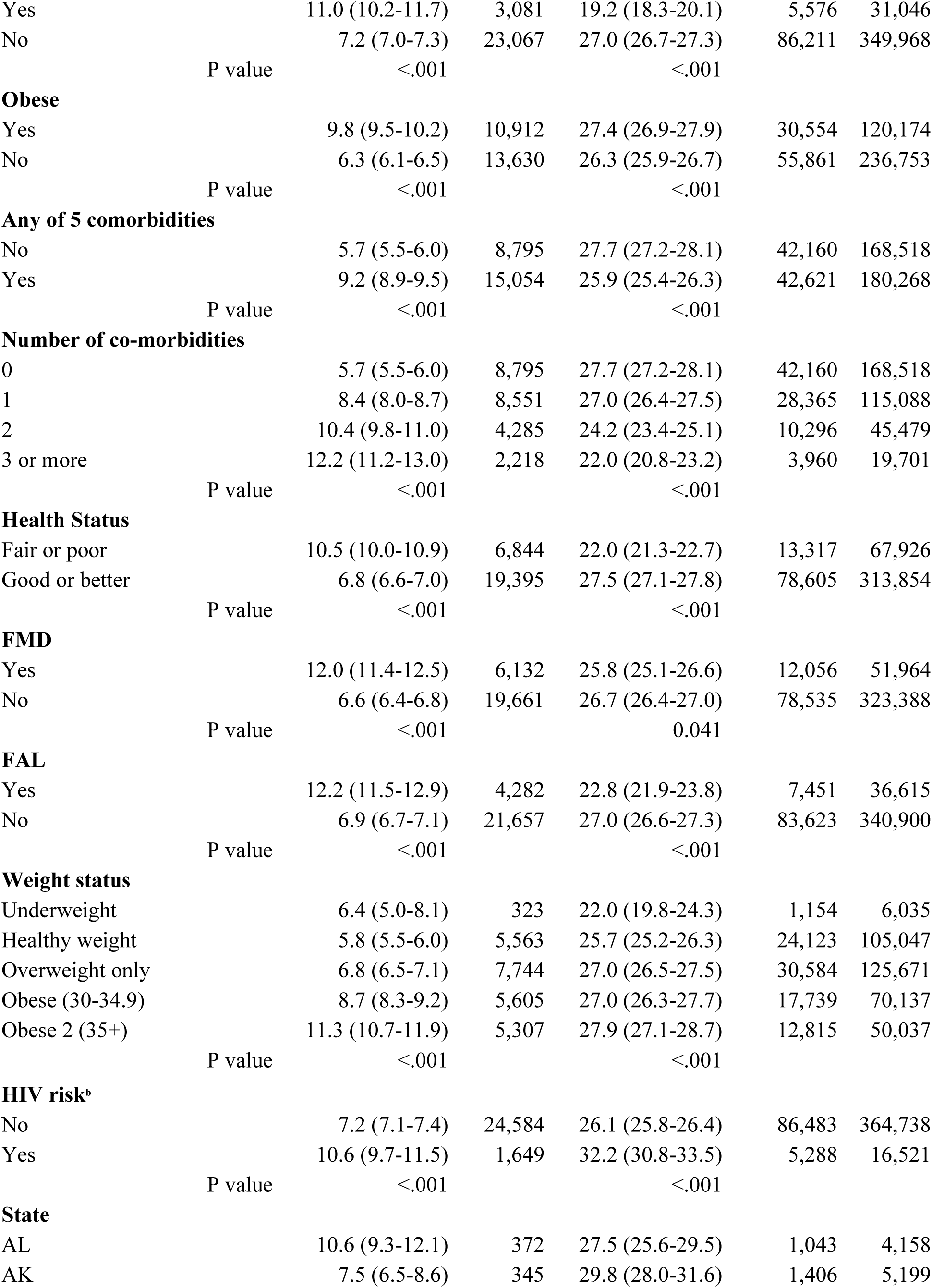
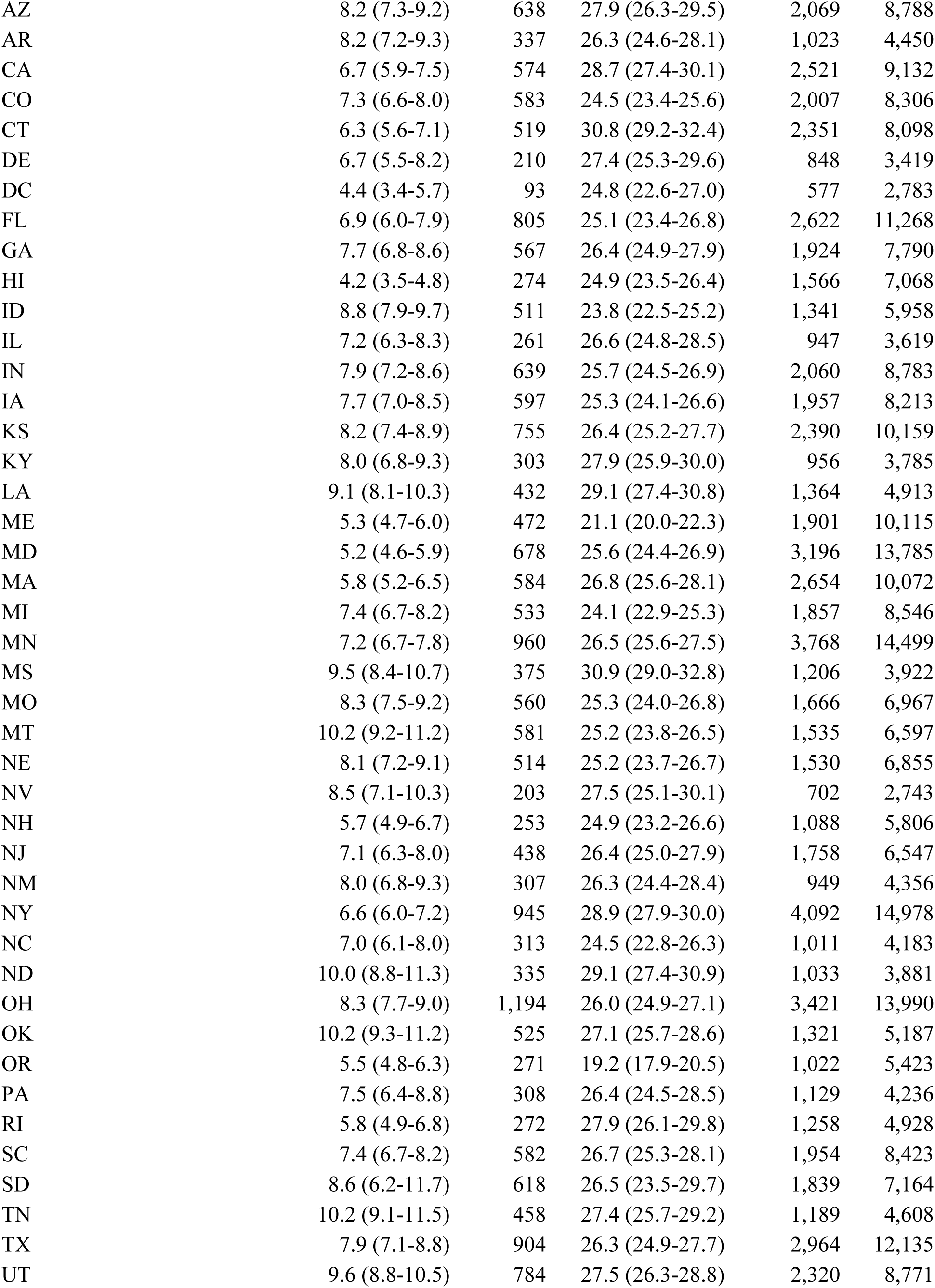
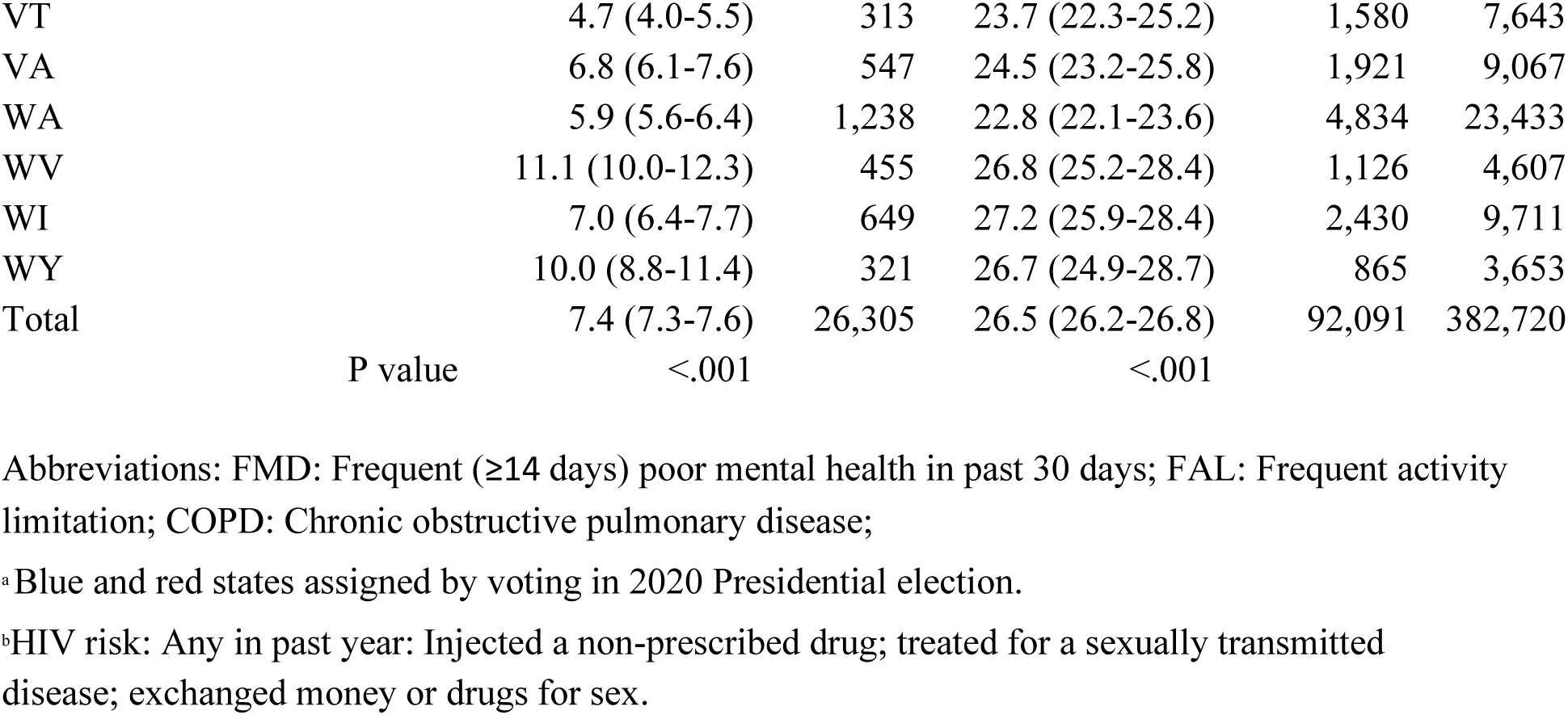
Long COVID (≥3 months) and just COVID (positive test only), 2022 Behavioral Risk Factor Surveillance System, 50 states plus DC. N= 382,720. Percents are weighted percentages and n’s are unweighted. Comorbidities: obesity, diabetes, cardiovascular disease (CVD), asthma, or chronic obstructive pulmonary disease (COPD).

Analysis of vaccination data from 29 states: Optional data on receipt of COVID vaccines were available [5] from a total of 29 states which included 5 states that asked the questions on different survey versions. Data from all survey versions were combined and re-weighted as described [5] under Complex Sampling Weights and Preparing Module Data for Analysis. The 29 states are: AR, CT, DE, GA, HI, ID, IL, IA, KS, LA, ME, MD, MA, MT, NE, NH, NJ, NM, NY, NC, ND, OK, RI, SC, TN, TX, WV, WI, and WY; 14 are blue states (N=100,179) and 15 are red states (N=102,023). Vaccine measures included receipt of any COVID vaccine, number of doses, and ≥3 doses vs <3 for 178,949 respondents.

## Results

As published earlier [4], 34.4% of respondents reported ever having a positive test for COVID, which included the new result of 26.5% (95% CI 26.2-26.8) with just COVID, ranging from 19.2% in OR to 30.9% in MS (Table 1). With 2,897 (0.75%) missing responses removed, 7.4% (7.3-7.6) had long COVID with rates ranging across states from 4.4% in DC to 11.1% in WV (Table 1). Results for the primary symptom experienced for long COVID were reported earlier [3, 4] with only 5.1% of adults with long COVID reporting no long-term symptoms that limited their activities [4]. Higher rates for just COVID (>30%) were found for ages 18-44 years, income ≥$75,000, adults with children in the household, current e-cigarette users, and reporting HIV risk (Table 1). Higher rates for long COVID were reported by these same groups except higher income, plus Hispanics and those with any of the 5 chronic conditions associated with COVID morbidity (obesity, asthma, CVD, diabetes, and COPD) [6,7] reported by 49.1% of all adults. Among adults with none of the 5 conditions, 5.7% reported long COVID compared with 12.1% of those with ≥3. Results for both outcomes were confirmed by logistic regression using models including all the measures shown (Table 2). With just COVID as the outcome the highest AOR was 2.01 for age 18-24 vs. age 65+ while the highest AOR for long COVID was 2.94 for respondents with ≥3 of the 5 chronic conditions vs. none. Results of comparisons using the 3-level measure found that that those with just a positive test had the best results for 15 of the 17 measures of chronic disease, disability, and health status among the 3 groups (Table 3), while those with long COVID had the worst results for 13 of the 17 measures.

**Table 2.** Weighted results of logistic regression, controlled for the measures listed, 2022 Behavioral Risk Factor Surveillance System, 50 states & DC, N=329,521. COVID risks=obesity, diabetes, cardiovascular disease, chronic obstructive pulmonary disease, asthma.

| **Outcome>>>**<br>**Group** | **Long COVID ≥3 months** |  | **Just COVID + test** |  |
| --- | --- | --- | --- | --- |
|  | **AOR (95% CI)** | **P value** | **AOR (95% CI)** | **P value** |
| Females w/kids v M, no kids | 2.21 (2.03-2.40) | <.001 | 1.12 (1.06-1.18) | <.001 |
| Females no kids v M no kids | 1.77 (1.65-1.89) | <.001 | 1.01 (0.97-1.05) | <.001 |
| Males w/kids v M no kids | 1.19 (1.08-1.31) | <.001 | 1.11 (1.05-1.17) | <.001 |
| 55-64 years v 65+ | 1.77 (1.62-1.94) | <.001 | 1.35 (1.28-1.43) | <.001 |
| 45-54 years v 65+ | 2.23 (2.02-2.46) | <.001 | 1.55 (1.46-1.64) | <.001 |
| 35-44 years v 65+ | 2.37 (2.13-2.64) | <.001 | 1.70 (1.60-1.81) | <.001 |
| 25-34 years v 65+ | 2.28 (2.06-2.53) | <.001 | 1.91 (1.80-2.02) | <.001 |
| 18-24 years v 65+ | 2.07 (1.84-2.34) | <.001 | 2.01 (1.88-2.15) | <.001 |
| Black v non-Hispanic white | 0.73 (0.67-0.80) | <.001 | 0.97 (0.92-1.03) | 0.289 |
| Hispanic v non-Hispanic white | 1.21 (1.04-1.41) | 0.016 | 0.90 (0.82-1.00) | 0.054 |
| Am. Indian v non-Hispanic white | 1.16 (0.94-1.42) | 0.158 | 0.91 (0.80-1.04) | 0.155 |
| Asian v. non-Hispanic white | 0.52 (0.42-0.65) | <.001 | 0.96 (0.88-1.06) | 0.445 |
| Other v. non-Hispanic white | 1.03 (0.94-1.13) | 0.522 | 1.10 (1.03-1.16) | 0.002 |
| \$25-<\$50K v < \$25K | 1.10 (0.99-1.22) | 0.067 | 1.26 (1.18-1.35) | <.001 |
| \$50-<\$75K v < \$25K | 1.18 (1.06-1.32) | 0.002 | 1.44 (1.34-1.54) | <.001 |
| \$75K-<\$100K v < \$25K | 1.23 (1.09-1.38) | 0.001 | 1.55 (1.44-1.67) | <.001 |
| \$100K+ v <\$25K | 1.04 (0.93-1.16) | 0.498 | 1.75 (1.64-1.88) | <.001 |
| Unknown v <\$25K income | 0.86 (0.77-0.97) | 0.012 | 1.15 (1.07-1.23) | <.001 |
| Annual flu shot v no | 0.86 (0.81-0.91) | <.001 | 0.97 (0.93-1.00) | 0.075 |
| Smoker v non-smoker | 0.77 (0.70-0.83) | <.001 | 0.67 (0.64-0.71) | <.001 |
| 1 COVID risk v 0 | 1.52 (1.42-1.62) | <.001 | 1.04 (1.00-1.08) | 0.043 |
| 2 COVID risk v 0 | 2.25 (2.07-2.44) | <.001 | 1.06 (1.00-1.12) | 0.033 |
| 3+COVID risk v 0 | 2.94 (2.64-3.27) | <.001 | 1.11 (1.03-1.20) | 0.009 |
| HIV risk<sup>a</sup> v no | 1.32 (1.19-1.47) | <.001 | 1.17 (1.09-1.25) | <.001 |
| E-cigarettes v no | 1.23 (1.12-1.36) | <.001 | 1.13 (1.06-1.21) | <.001 |
| Red states v Blue states<sup>b</sup> | 1.15 (1.09-1.22) | <.001 | 1.00 (0.96-1.03) | 0.891 |
| **COVID risks if entered separately** |  |  |  |  |
| Obesity | 1.41 (1.33-1.50) | <.001 | 1.07 (1.04-1.11) | <.001 |
| Diabetes | 1.22 (1.12-1.33) | <.001 | 1.08 (1.02-1.14) | 0.008 |
| Cardiovascular disease | 1.30 (1.19-1.42) | <.001 | 0.99 (0.93-1.05) | 0.666 |
| COPD | 1.56 (1.40-1.73) | <.001 | 0.89 (0.83-0.96) | 0.002 |
| Asthma | 1.63 (1.51-1.77) | <.001 | 1.02 (0.96-1.08) | 0.488 |

Abbreviations: AOR: adjusted odds ratio; COPD: chronic obstructive pulmonary disease;
<sup>a</sup>HIV risk: Any in past year: Injected a non-prescribed drug; treated for a sexually transmitted disease; exchanged money or drugs for sex.
<sup>b</sup> Assigned by voting in 2020 Presidential election.

**Table 3.** Direct comparison of measures of long COVID^a^, just a positive test^b^, and neither 2022 Behavioral Risk Factor Surveillance System, all age 18+, N=382,720.

| % of this measure among these adults><br>**Health conditions** | Long<br>COVID<sup>a</sup> | + test<sup>b</sup> | Neither | Total | P value |
| --- | --- | --- | --- | --- | --- |
| **N=** | 26,305 | 92,091 | 264,324 | 382,720 |  |
| Any CVD | 9.8 | **7.1** | 10.2 | 9.3 | <.001 |
| 95% CIs | 9.2-10.5 | 6.8-7.5 | 9.9-10.4 | 9.2-9.5 |  |
| Arthritis | 33.2 | **23.3** | 28.2 | 27.3 | <.001 |
| 95% CIs | 32.1-34.3 | 22.8-23.9 | 27.9-28.6 | 27.0-27.6 |  |
| Diabetes | 13.2 | **10.7** | 12.5 | 12.1 | <.001 |
| 95% CIs | 12.4-14.0 | 10.3-11.1 | 12.2-12.8 | 11.9-12.3 |  |
| COPD | 10.1 | **5.0** | 7.2 | 6.9 | <.001 |
| 95% CIs | 9.4-10.9 | 4.7-5.2 | 7.1-7.4 | 6.7-7.0 |  |
| Asthma (current) | 17.6 | 9.9 | 9.1 | 10.0 | <.001 |
| 95% CIs | 16.7-18.5 | 9.5-10.3 | 8.9-9.3 | 9.8-10.1 |  |
| Cancer other than skin includes<br>melanoma | 7.9 | **7.0** | 9.0 | 8.4 | <.001 |
| 95% CIs | 7.3-8.5 | 6.7-7.3 | 8.8-9.2 | 8.2-8.6 |  |
| **Disability and related issues** |  |  |  |  |  |
| Deaf or difficulty hearing | 7.6 | **5.5** | 7.6 | 7.1 | <.001 |
| 95% CIs | 7.0-8.3 | 5.2-5.8 | 7.4-7.9 | 6.9-7.2 |  |
| Blind or serious problems seeing | 6.7 | **4.5** | 5.9 | 5.6 | <.001 |
| 95% CIs | 6.1-7.2 | 4.2-4.9 | 5.7-6.1 | 5.4-5.8 |  |
| Difficulty walking/climbing stairs | 17.5 | **9.9** | 14.9 | 13.7 | <.001 |
| 95% CIs | 16.6-18.4 | 9.5-10.3 | 14.6-15.2 | 13.5-14.0 |  |
| Difficulty bathing/dressing | 5.3 | **2.6** | 4.1 | 3.8 | <.001 |
| 95% CIs | 4.8-5.8 | 2.4-2.9 | 3.9-4.2 | 3.6-3.9 |  |
| Difficulty doing errands alone | 11.2 | **5.8** | 8.4 | 7.9 | <.001 |
| 95% CIs | 10.5-12.1 | 5.5-6.1 | 8.2-8.7 | 7.7-8.1 |  |
| Difficulty with memory, concentrating | 21.9 | **11.9** | 13.1 | 13.5 | <.001 |
| 95% CIs | 20.9-22.9 | 11.5-12.4 | 12.8-13.4 | 13.2-13.7 |  |
| **Health status & related** |  |  |  |  |  |
| Fair or poor health status | 25.2 | **14.9** | 18.3 | 17.9 | <.001 |
| 95% CIs | 24.2-26.3 | 14.4-15.4 | 18.0-18.7 | 17.7-18.2 |  |
| Frequent mental distress (FMD) | 25.6 | 15.5 | 15.0 | 15.9 | <.001 |
| 95% CIs | 24.6-26.7 | 15.0-16.0 | 14.7-15.3 | 15.7-16.2 |  |
| Frequent physical distress (FPD) | 20.7 | **10.8** | 12.5 | 12.7 | <.001 |
| 95% CIs | 19.6-21.7 | 10.4-11.3 | 12.2-12.8 | 12.4-12.9 |  |
| Frequent activity limitation (FAL) | 16.2 | **8.5** | 9.7 | 9.9 | <.001 |
| 95% CIs | 15.3-17.1 | 8.1-8.9 | 9.5-10.0 | 9.7-10.1 |  |
| Unable to work | 8.1 | **4.6** | 7.1 | 6.5 | <.001 |
| 95% CIs | 7.5-8.8 | 4.3-4.9 | 6.9-7.4 | 6.4-6.7 |  |
| **Health behaviors and risk factors** |  |  |  |  |  |
| All overweight (BMI>25) | 74.9 | 69.1 | 66.3 | 67.7 | <.001 |
| 95% CIs | 73.9-76.0 | 68.4-69.7 | 65.9-66.7 | 67.4-68.0 |  |
| Obese (BMI=30+) | 44.1 | 34.5 | 32.0 | 33.6 | <.001 |
| 95% CIs | 42.9-45.4 | 33.9-35.2 | 31.6-32.4 | 33.3-33.9 |  |
| Current smoking | 12.3 | **9.5** | 13.9 | 12.6 | <.001 |
| 95% CIs | 11.6-13.1 | 9.1-9.9 | 13.6-14.2 | 12.4-12.9 |  |
| Heavy drinking | 7.1 | 6.9 | 6.4 | 6.6 | 0.006 |
| 95% CIs | 6.6-7.8 | 6.5-7.2 | 6.2-6.6 | 6.4-6.7 |  |
| E-cigarettes current | 10.0 | 8.7 | 6.6 | 7.4 | <.001 |
| 95% CIs | 9.3-10.7 | 8.4-9.1 | 6.4-6.9 | 7.3-7.6 |  |
| Any 5 co-morbidities/risk factors <sup>c</sup> | 61.0 | 47.7 | 48.8 | 49.4 | <.001 |
| 95% CIs | 59.7-62.2 | 47.0-48.4 | 48.3-49.2 | 49.0-49.7 |  |
| No leisure time activity (sedentary) | 24.9 | **20.6** | 24.5 | 23.5 | <.001 |
| 95% CIs | 23.9-26.0 | 20.0-21.1 | 24.1-24.8 | 23.2-23.8 |  |
| Depression diagnosis | 33.9 | 20.7 | 20.0 | 21.2 | <.001 |
| 95% CIs | 32.7-35.0 | 20.2-21.2 | 19.7-20.4 | 21.0-21.5 |  |
| **Health access measures** |  |  |  |  |  |
| No health care coverage | 8.3 | **6.6** | 9.0 | 8.3 | <.001 |
| 95% CIs | 7.6-9.1 | 6.2-7.0 | 8.7-9.3 | 8.1-8.5 |  |
| Medicaid coverage | 10.6 | 7.9 | 8.5 | 8.5 | <.001 |
| 95% CIs | 9.9-11.5 | 7.5-8.3 | 8.3-8.8 | 8.3-8.7 |  |
| Unable to see MD because of cost | 19.4 | 9.9 | 10.6 | 11.0 | <.001 |
| 95% CIs | 18.3-20.4 | 9.5-10.3 | 10.3-10.8 | 10.8-11.3 |  |
| **Other** |  |  |  |  |  |
| Females | 63.4 | 50.6 | 50.0 | 51.1 | <.001 |
| 95% CIs | 62.3-64.6 | 49.9-51.2 | 49.5-50.4 | 50.8-51.5 |  |
| Ages 18-64 | 86.3 | 83.8 | 72.4 | 76.5 | <.001 |
| 95% CIs | 85.5-87.1 | 83.3-84.2 | 72.1-72.7 | 76.2-76.7 |  |
| ≥3 vaccines <sup>d</sup> | 35.6 | 42.7 | 50.3 | 47.1 | <.001 |
| 95% CIs | 33.9-37.3 | 41.7-43.6 | 49.6-50.9 | 46.6-47.6 |  |

<sup>a</sup> Long COVID: symptoms lasting ≥3 months
<sup>b</sup> Just a positive COVID test, not long COVID
<sup>c</sup> Obesity, cardiovascular disease, chronic obstructive pulmonary disease, diabetes or asthma.
<sup>d</sup> 29 states that asked optional vaccine questions; AR, CT, DE, GA, HI, ID, IL, IA, KS, LA, ME, MD, MA, MT, NE, NH, NJ, NM, NY, NC, ND, OK, RI, SC, TN, TX, WV, WI, and WY.
Figures in bold represent lowest among the 4 groups,

## Vaccinations

Vaccination data from the 29 states showed that associations with COVID outcomes were similar for adults with 0, 1, or 2 vaccines as were results for 3 or 4+ vaccines so the measure of ≥3 vaccine doses vs <3 was chosen for most analyses. Among adults age 65+, 67.8% reported ≥3 vaccines compared with 40.4% for adults 18-64 years and 46.9% for all respondents with non-missing age. For all adults, those with long COVID were the least likely of the 3 COVID groups to report ≥3 vaccines; 35.6% for adults with long COVID, 42.7% for just a positive test, 50.3% for those never testing positive and 47.1% overall. Long COVID rates were slightly (but not significantly) higher in the 29 states vs. all 50 at 7.5% (7.3-7.8) and ≥3vaccinations appeared to reduce long COVID rates by 38% (from 9.2% to 5.7%) and just COVID rates by 16.2% (from 29.0% to 24.3%). Apparent effectiveness of ≥3 vaccines was confirmed in logistic regression with AORs of 0.66 for long COVID and 0.77 for just COVID (Table S-1).

Results for all 50 states (Tables 1 & 2) showed that red states had significantly higher long COVID rates and AORs than blue states while red and blue states had similar rates for just COVID. Census regions and red/blue states overlapped to some extent: >70% of states in the Northeast or West were blue states while >70% of states in the Midwest or South were red states (not shown). Significant differences between red and blue states were also found for 4 of the 5 risk factors: diabetes (13.0% in red states vs. 11.3% in blue states), obesity (35.8% vs. 31.4%), CVD (10.2% vs. 8.5%), and COPD (7.9% vs. 6.0%), but not asthma The percentage reporting ≥3 COVID vaccines from the 29 states was also different for red (40.1%) vs. blue states (53.3%). Apparent vaccine effectiveness of ≥3 vs. <3 vaccines at reducing long COVID rates was similar in red and blue states at 35% in red states (reduction of rate from 9.9% to 6.4%) and 37% in blue states (from 8.4% to 5.3%).

## Discussion

This study used 2022 population-based data to compare adults with long COVID, those with just a positive test, and neither, finding that adults who reported just COVID were very different from adults reporting long COVID and rates varied widely across states. Those with just COVID had the lowest (best) rates for 15 of 17 separate measures of chronic conditions, disability, and health status among the 3 groups being compared (Table 3) while adults with long COVID had the worst rates for 13 of those same 17 measures. Risk factor results were also quite different for just COVID and long COVID. In unadjusted and adjusted results, increasing numbers of the risk factors of obesity, diabetes, COPD, CVD, and asthma identified earlier [6,7] increased rates of long COVID from 5.7% for zero risk factors to 12.2% for 3 or more, with ≥3 risk factors having the highest AOR of 2.93. For just COVID, unadjusted results in Table 1 showed an inverse association with increasing numbers of the 5 risk factors from a rate of 27.7% for 0 risk factors down to 22.0% for ≥3, while adjusted results found younger age and higher income had highest AORs. Because both long COVID and just COVID were more prevalent among younger adults, 86.3% of adults with long COVID and 83.8% of those with just a positive test were ages 18-64 years. Long COVID, but not just COVID, had significantly higher rates in red vs. blue states which remained when results were controlled for all the measures included in logistic regression. Vaccine results, although limited because only 29 states provided data, showed that ≥3 vaccines reduced long COVID rates by 38% vs. 16.2 % for just a positive test. The role of obesity in COVID risk appears to remain when the outcome is either long COVID or just COVID instead of deaths [8], as seen in Tables 1 and 2. In addition to being a risk factor for COVID complications, obesity has been shown to have a negative impact on immune response [9,10].

Remembering pandemic history might help to put COVID and long COVID in perspective. Before vaccines were available, hospitals and health care workers were overwhelmed and death rates, especially among older adults were high. The situation has changed since vaccines became available in 2021, but the total number of US deaths had reached over one million by the end of 2022 [11]. Results from this study indicate that 34.4% of adults surveyed in 2022 reported they had ever had a positive test, representing 89.8 million of the 261 million adults. Therefore, a rough estimate of case fatality rate (deaths/cases) is about 1%. Those deaths include 415,399 COVID deaths in 2021 and 244,986 deaths in 2022 [12] indicating that COVID deaths are decreasing over time. Long COVID was reported by one-fifth of all respondents reporting a positive test, with a wide range across states, or an estimated 19.3 million adults, and about 69.2 million had only a positive test. While not necessarily fatal (we can’t tell from these data), long COVID appears to be serious and chronic (by definition, lasting ≥3 months). Among adults with long COVID in this study, 95% reported symptoms which limited their activities. As of 2022, there were an estimated 69.2 million adults with just a positive COVID test that did not develop into long COVID, but these adults were more likely to report better health status compared with other survey respondents.

How can the above information be used to reduce future impact of COVID and especially long COVID? The decrease in deaths between 2021 and 2022 is likely related to increased immunity to COVID from a combination of natural immunity from the approximately 90 million adults who reported a positive test and from vaccines. Higher vaccination rates among adults 65+ may help account for their lower rates of long COVID and just a positive test compared with younger adults, as found in this study and a previous study [3]. These study results also indicate that one in 5 (over a wide range among states) of those who had a positive COVID test reported long COVID that lasted ≥3 months and caused activity limitations to most. Because common risk factors including obesity and diabetes were shown to increase the risk (odds) of developing long COVID, a typical strategy might be interventions to lower the prevalence of risk factors. But success of such interventions is rare and indeed, risk factors such as obesity [13] and diabetes have been increasing steadily over time. Our finding that long COVID prevalence was reduced 38% (from 9.2% to 5.7%) among adults reporting ≥3 vaccine doses vs. those with <3 doses suggests that increasing vaccination rates could be an effective strategy. According to results in Table 1. in order to reduce long COVID rates down to approximately 5.7% by targeting the 5 risk factors would require reducing the number of risk factors to zero! However, due to variation in long COVID rates the 5.7% rate is unlikely to be the same in all states and increasing vaccination rates still appears more feasible than trying to reduce risk factors. Further study of vaccines and COVID for all 50 states is also recommended. Vaccine promotions might need to be carefully tailored to motivate people most likely to resist vaccination [14] and targeted to adults with risk factors, but still seem the best choice.

## Red and blue state comparisons

Some reminders about the red state/blue state data; although the variable defining red and blue states is based on voting in the 2020 Presidential election it should be clear that there are other differences between the states that impact COVID. Adults in red states are significantly more likely to report 4 of the 5 risk factors for long COVID – obesity, diabetes, CVD, and COPD – compared to those in blue states. Although limited to 29 states, the vaccination data also clearly show that adults in red states are less likely to be vaccinated for COVID. Other study results indicate that number of risk factors and vaccination rates are associated with long COVID rates, so it should not be surprising that red states have higher long COVID rates. These appear to be factors that can be modified by behavior change, with the potential for reducing long COVD rates in red states.

## Comparisons with other studies

Other studies using different sources of data have found different rates of long COVID but this rate of 7.4% from the BRFSS is consistent with the 6% recently reported from another survey with the same definition of long COVID [15] but is much higher than the 1.7% in another study where long COVID was defined as symptoms lasting 2 months [16].That result is not inconsistent with the wide range of rates this study found among states and could depend on the definition used for long COVID, when results were obtained, and which variants were involved. We found no other studies comparing long COVID with just having a positive test for COVID. The study result for activity limitation among adults with long COVID of 95% is considerably higher than the results of another recent survey where the figure was 26% for “significant” activity limitation [16]. That difference might be due to the adjective “significant” only in the other survey. Further study is needed but these results already suggest a need for support services for people with long COVID.

Vaccine studies have found varying results, depending on the outcome studied, the year, and the variants involved. As one meta-analysis with long COVID as outcome [17] notes, the effectiveness of COVID vaccines on long COVID varies. Two-doses of vaccine lowered risk of COVID compared to no vaccination (OR, 0.64; 95% CI 0.45-0.92) while a single dose did not appear effective. On the other hand, this study suggests that ≥3 vaccine doses are needed to be effective, and we found results to be similar for 0, 1, or 2 doses which was our comparison group. A study in Australia [18] also found 3 doses of vaccine were needed to be effective against the Omicron variant of COVID.

Limitations: There are at least five limitations to this study. First, because the BRFSS only surveys households, among the institutions the survey omits are nursing homes and prisons which appeared to have high rates of COVID especially early in the pandemic. Thus, results may underestimate the true rates of COVID and long COVID. Second, results are self-reported and except as noted for COVID are not based on an actual test or diagnosis; the implications of this limitation are unknown. Third, the lack of a measure of hypertension on the survey for 2022 meant that the composite measure of comorbidities lacked a key component (6,7). Fourth, survey results can’t distinguish cause and effect so results indicating higher rates of measures such as cognitive disability and frequent activity limitation only indicate an association and could represent risk factors and/or consequences of long COVID. Fifth, vaccination data are only available for 29 states. Lastly, while long COVID was defined as lasting ≥3 months the survey did not determine whether symptoms were temporary or lasted beyond 3 months or whether hospitalizations (or deaths) were involved. The study left many questions about long COVID unanswered and suggesting further investigation.

Strengths: The study also benefitted from using population-based data from all 50 states and DC that were weighted to reflect the population of each state by demographic groups. More importantly, prevalence rates of COVID measures reported for each state showed the wide range of values across states.

## Conclusion

This study shows that results for just a COVID positive test and long COVID are quite different and each varied widely across states. First, the former compared favorably with both other groups on health and disability measures, while those with long COVID reported consistently poorer health status, disability, and chronic conditions and 95% reported activity limitations. Second, both a positive test and long COVID rates were higher among adults ages 18-64 compared with those 65 and older, contrary to earlier results based on death data. Third, adjusted results showed that more of the risk factors that were found to increase risk of hospitalization for COVID (obesity, asthma, CVD, diabetes, and COPD)[6,7] also had the highest AORs for long COVID which affected one-fifth of adults with a positive COVID test. For the other 80% with a positive test (the just COVID group), younger age and higher income had the greatest risk, but results suggest this group is in relatively good health. Fourth, regional and red vs. blue state differences were seen for long COVID but not for just COVID rates, which may suggest the need to address regions and states differently. These differences may also suggest realistic goals to set. Fifth, limited data for vaccines show the effectiveness of ≥3 doses at reducing rates of COVID and especially long COVID rates. Effectives of vaccines at reducing long COVID rates appears similar in red and blue states. We conclude that the most effective way to address COVID now is to concentrate on long COVID with targeted vaccine promotions for adults with risk factors.

## Data Availability

Data used are available at https://www.cdc.gov/brfss/annual_data/annual_2022.html.

## Acknowledgment

This study was partially supported by a cooperative agreement from CDC (#1NU58DP006901) which had no role in the writing of the manuscript.

The authors have no conflicts of interest to disclose.

## References

[1]. CDC. Long COVID or Post-COVID Conditions. 2023. Available online: https://www.cdc.gov/coronavirus/2019-ncov/long-term-effects/index.html (accessed on 27 January 2024).

[2]. Pierannunzi C, Hu SS, Balluz L. A systematic review of publications assessing reliability and validity of the Behavioral Risk Factor Surveillance System (BRFSS), 2004-2011. BMC Med Res Methodol. 2013;13:49.

[3]. Nguyen, K.H.; Bao, Y.; Mortazavi, J.; Allen, J.D.; Chocano-Bedoya, P.O.; Corlin, L. Prevalence and Factors Associated with Long COVID Symptoms among U.S. Adults, 2022. Vaccines 2024, 12, 99. 10.3390/vaccines12010099.

[4]. Adams ML, Grandpre J. Prevalence of COVID-19 and long COVID – Results from the 2022 Behavioral Risk Factor Surveillance System, 50 states. MedRxiv preprint doi: 10.1101/2023.09.29.23296352; posted September 29, 2023.

[5]. Behavioral Risk Factor Surveillance System (BRFSS). Centers for Disease Control and Prevention. Behavioral Risk Factor Surveillance System. 2022 Survey data and documentation. https://www.cdc.gov/brfss/annual_data/annual_2022.html. Accessed September 1, 2023.

[6]. Garg S, Kim L, Whitaker M, O’Halloran A, Cummings C, Holstein R, et al. Hospitalization rates and characteristics of patients hospitalized with laboratory-confirmed coronavirus disease 2019—COVID-NET, 14 States, March 1–30, 2020. MMWR Morb Mortal Wkly Rep. 2020;69:458–64.

[7]. Adams, M. L., Katz, D. L., & Grandpre, J. (2020). Updated Estimates of Chronic Conditions Affecting Risk for Complications from Coronavirus Disease, United States. Emerging Infectious Diseases, 2020 Sep;26(9):2172–2175. doi: 10.3201/eid2609.202117.

[8]. Adams, M. L. (2022). Accounting for the Origins and Toll of COVID 19 The key role of Overweight in COVID-19. Am J Health Promot, Feb;36(2), 385–387.

[9]. Amin MT, Fatema K, Arefin S, Hussain F, Bhowmik DR, Hossain MS. Obesity, a major risk factor for immunity and severe outcomes of COVID-19. Biosci Rep. 2021 Aug 27;41(8):BSR20210979. doi: 10.1042/BSR20210979. PMID: 34350941; PMCID: PMC8380923.

[10]. Ealey KN, Phillips J, Sung HK. COVID-19 and obesity: fighting two pandemics with intermittent fasting. Trends Endocrinol Metab. 2021 Sep;32(9):706–720. doi: 10.1016/j.tem.2021.06.004. Epub 2021 Jun 25. PMID: 34275726; PMCID: PMC8226104.

[11]. USA Facts. US COVID-19 cases and deaths by state through date specified (December 31, 2022) https://usafacts.org/visualizations/coronavirus-covid-19-spread-map. Accessed 12/31/2022.

[12]. Ahmad FB, Cisewski JA, Xu J, Anderson RN. Provisional Mortality Data — United States, 2022. MMWR Morb Mortal Wkly Rep 2023;72:488–492. DOI: 10.15585/mmwr.mm7218a3).

[13]. Pernenkil V, Wyatt T, Akinyemiju T. Trends in smoking and obesity among US adults before, during, and after the great recession and Affordable Care Act roll-out. Prev Med. 2017 Sep; 102:86–92. doi:10.1016/j.ypmed.2017.07.001. Epub 2017 Jul 8.

[14]. Robertson CT, Bentele K, Meyerson B, et al. Effects of political versus expert messaging on vaccination intentions of Trump voters. PLoS One 16 (2021): e0257988.

[15]. Ford ND, Slaughter D, Edwards D, et al. Long COVID and Significant Activity Limitation Among Adults, by Age — United States, June 1–13, 2022, to June 7–19, 2023. MMWR Morb Mortal Wkly Rep 2023;72:866–870. DOI: 10.15585/mmwr.mm7232a3

[16]. Perlis RH, Santillana M, Ognyanova K, Safarpour A, Lunz Trujillo K, Simonson MD, Green J, Quintana A, Druckman J, Baum MA, Lazer D. Prevalence and Correlates of Long COVID Symptoms Among US Adults. JAMA Netw Open. 2022 Oct 3;5(10):e2238804. doi: 10.1001/jamanetworkopen.2022.38804.

[17]. Watanabe A, Iwagami M, Yasuhara J, Takagi H, Kuno T. Protective effect of COVID-19 vaccination against long COVID syndrome: A systematic review and meta-analysis. Vaccine. 2023 Mar 10;41(11):1783–1790. doi: 10.1016/j.vaccine.2023.02.008. Epub 2023 Feb 8. PMID: 36774332; PMCID: PMC9905096.

[18]. Liu B, Gidding H, Stepien S, Cretikos M, Macartney K. Relative effectiveness of COVID-19 vaccination with 3 compared to 2 doses against SARS-CoV-2 B.1.1.529 (Omicron) among an Australian population with low prior rates of SARS-CoV-2 infection. Vaccine. 2022 Oct 12;40(43):6288–6294. doi: 10.1016/j.vaccine.2022.09.029. Epub 2022 Sep 21. PMID: 36180375; PMCID: PMC9489983.

