## Supplementary material for "Comparing surveyed adults with long COVID and those with just a positive test helps put COVID into perspective": Table S-1

| Table S-1. Weighted results of logistic regression with vaccine data, controlled for the measures listed, 2022 Behavioral Risk Factor Surveillance System, 29 states, N=123,202 adults, age 18+. | | | | | | | | | | |
| --- | --- | --- | --- | --- | --- | --- | --- | --- | --- | --- |
| COVID risks=obesity, diabetes, cardiovascular disease, chronic obstructive pulmonary disease, asthma. | | | | | | | | | | |
| Adjusted odds ratios (AOR) and 95% confidence intervals | | | | | | |  | |  | |
|  | Long COVID (≥3 months) | | | | | Just Positive test | | | | |
| **Group** | **AOR** | **95% CI** | | **P>t** | | **AOR** | | **95% CI** | | **P>t** |
| Females w/kids v M, no kids | 2.02 | 1.77-2.30 | | <.001 | | 1.20 | | 1.10-1.30 | | <.001 |
| Females no kids v M no kids | 1.66 | 1.48-1.87 | | <.001 | | 1.02 | | 0.95-1.09 | | 0.631 |
| Males w/kids v M no kids | 0.97 | 0.83-1.13 | | 0.673 | | 1.17 | | 1.07-1.27 | | <.001 |
| 55-64 years v 65+ | 1.62 | 1.41-1.85 | | <.001 | | 1.29 | | 1.18-1.41 | | <.001 |
| 45-54 years v 65+ | 2.17 | 1.87-2.51 | | <.001 | | 1.41 | | 1.28-1.55 | | <.001 |
| 35-44 years v 65+ | 2.12 | 1.81-2.47 | | <.001 | | 1.53 | | 1.38-1.69 | | <.001 |
| 25-34 years v 65+ | 2.12 | 1.81-2.48 | | <.001 | | 1.71 | | 1.55-1.89 | | <.001 |
| 18-24 years v 65+ | 1.78 | 1.45-2.18 | | <.001 | | 1.83 | | 1.62-2.06 | | <.001 |
| Black v non-Hispanic white | 0.75 | 0.65-0.88 | | <.001 | | 0.97 | | 0.89-1.06 | | 0.538 |
| Hispanic v non-Hispanic white | 0.97 | 0.77-1.23 | | 0.82 | | 1.01 | | 0.84-1.22 | | 0.88 |
| Am. Indian v non-Hispanic white | 1.00 | 0.76-1.32 | | 0.976 | | 0.99 | | 0.79-1.23 | | 0.913 |
| Asian v. non-Hispanic white | 0.55 | 0.40-0.76 | | <.001 | | 0.95 | | 0.80-1.13 | | 0.579 |
| Other v. non-Hispanic white | 1.02 | 0.88-1.18 | | 0.772 | | 1.03 | | 0.94-1.13 | | 0.517 |
| $25-<$50K v < $25K | 1.26 | 1.07-1.49 | | 0.006 | | 1.33 | | 1.20-1.48 | | <.001 |
| $50-<$75K v < $25K | 1.28 | 1.08-1.51 | | 0.005 | | 1.51 | | 1.35-1.69 | | <.001 |
| $75K-<$100K v < $25K | 1.42 | 1.18-1.70 | | <.001 | | 1.55 | | 1.38-1.74 | | <.001 |
| $100K+ v <$25K | 1.20 | 1.01-1.42 | | 0.041 | | 1.82 | | 1.64-2.02 | | <.001 |
| Unknown v <$25K income | 0.95 | 0.79-1.14 | | 0.602 | | 1.19 | | 1.06-1.33 | | 0.004 |
| Annual flu shot v no | 0.94 | 0.85-1.04 | | 0.218 | | 1.11 | | 1.04-1.18 | | 0.001 |
| Smoker v non-smoker | 0.77 | 0.66-0.89 | | 0.001 | | 0.70 | | 0.64-0.76 | | <.001 |
| 1 COVID risk v 0 | 1.50 | 1.36-1.66 | | <.001 | | 1.08 | | 1.01-1.15 | | 0.016 |
| 2 COVID risk v 0 | 2.13 | 1.87-2.43 | | <.001 | | 1.05 | | 0.96-1.15 | | 0.267 |
| 3+COVID risk v 0 | 3.22 | 2.72-3.82 | | <.001 | | 1.07 | | 0.93-1.22 | | 0.368 |
| HIV risk^a^ v no | 1.36 | 1.14-1.62 | | 0.001 | | 1.13 | | 1.01-1.27 | | 0.031 |
| E-cigarettes v no | 1.18 | 1.01-1.38 | | 0.034 | | 1.12 | | 1.01-1.24 | | 0.037 |
| Red states v Blue states^b^ | 1.09 | 1.00-1.19 | | 0.049 | | 0.95 | | 0.90-1.00 | | 0.053 |
| COVID vaccine ≥3 doses v <3 | 0.66 | 0.59-0.73 | | <.001 | | 0.77 | | 0.73-0.82 | | <.001 |
| ^a^HIV risk: Any in past year: Injected a non-prescribed drug; treated for a sexually transmitted | | | | | | | | | | |
| disease; exchanged money or drugs for sex, anal sex without a condom, or 4+ sex partners. | | | | | | | | | | |
| ^b^ Assigned by voting in 2020 Presidential election. | | | | |  | |  | |  | |
| 29 States are: AR, CT, DE, GA, HI, ID, IL, IA, KS, LA, ME, MD, MA, MT, NE, NH, NJ, NM, NY, NC, | | | | | | | | | | |
| ND, OK, RI, SC, TN, TX, WV, WI, and WY. | | |  | |  | |  | |  | |
